# Development of Colorimetric and Fluorometric Loop-Mediated Isothermal Amplification (LAMP) Assays for the Point-of-Care Molecular Diagnosis of *Blastocystis* spp

**DOI:** 10.1101/2025.10.24.25338741

**Authors:** Roaa Zedan, Huseyin Tombuloglu, Ayman Elbadry

## Abstract

*Blastocystis* species (spp.) is a genetically diverse protozoan parasite found in the digestive tract of animals and humans. Current clinical diagnosis relies heavily on wet mount microscopy, a method with known limitations, as well as molecular techniques like PCR, nested-PCR, and RT-PCR. However, neither microscopy nor PCR-based approaches are a perfect standard method due to their limitations, such as low accuracy, high equipment cost, and the need for well-trained personnel. Loop-mediated isothermal amplification (LAMP) technique offers the advantage of eliminating these limitations. Therefore, this study aimed to develop rapid and accurate fluorometric LAMP (fLAMP) and colorimetric LAMP (cLAMP) assays. We collected a total of 197 stool samples from both asymptomatic individuals and patients with gastrointestinal conditions. The performance of the LAMP assays was evaluated and compared against a standard RT-PCR assay. Results showed that both LAMP assays provided rapid detection of *Blastocystis* spp. in 20.0 to 29.2 minutes. The cLAMP assay demonstrated a sensitivity of 95.77%, a specificity of 85.00%, and an accuracy of 90.97%. The fLAMP assay exhibited a sensitivity of 91.43%, a specificity of 93.26%, and an accuracy of 92.27%. The limit-of-detection (LoD) of the fLAMP and cLAMP assays was 5×10^−6^ ng/µL, which corresponds to 5 fg/µL. The cLAMP assay offers a distinct advantage with its simplified visual result interpretation via a naked-eye color change, eliminating the need for complex instruments. This makes the cLAMP assay suitable for point-of-care testing (PoCT). These characteristics, combined with the femtogram-level detection capability of both assays, make the assays suitable for testing samples containing low DNA copy numbers. This is the first study to develop and elucidate the performance of two LAMP-based assays for the diagnosis of *Blastocystis* spp. The cLAMP assay exhibits potential to be used for PoCT, particularly in resource-limited settings. Their speed, accuracy, and analytical sensitivity hold promise for improving the diagnosis of *Blastocystis* spp. infections in patients with gastrointestinal symptoms.

## INTRODUCTION

*Blastocystis* spp. is a protozoan parasite that lives in the intestines of both humans and animals. This is the most common communal eukaryotic parasite found in human faecal specimens. Its transmission is likely to occur faeco-oral via contaminated food and drink (Estes, 2016; Moe et al., 1997; Yoshikawa et al., 2004). It is an extremely genetically diverse unicellular protozoan parasite with uncertain pathogenicity (Skotarczak, 2018). To date, 28 distinct subtypes (STs) of *Blastocystis* spp. have been identified in human hosts. This significant genetic diversity has led to the scientific consensus that the term “*Blastocystis* spp.” is the more accurate and appropriate designation for *Blastocystis* detected in humans, as it reflects the fact that multiple species or closely related genetic lineages within the *Blastocystis* genus can infect humans.

Among the various subtypes identified in humans, ST-1, ST-2, ST-3, and ST-4 are recognized as the most prevalent globally. However, the distribution of these and other subtypes can vary geographically and across different populations. The recognition of these subtypes is crucial for epidemiological studies, understanding potential differences in pathogenicity, and investigating transmission dynamics of *Blastocystis* infections. Other subtypes are sporadic and could potentially be linked to zoonotic transmission (Rudzińska & Sikorska, 2023). Human and a variety of non-human animal feces have been found to contain *Blastocystis* (e.g., canids, pigs, monkeys, rodents, birds, etc.). There are continuing efforts to describe the host distribution and geographic location of different *Blastocystis* subtypes (Rudzińska & Sikorska, 2023). It is still up for controversy and investigation as to whether *Blastocystis* spp. (or certain subtypes thereof, or specific strains of certain subtypes) may cause gastrointestinal illness in people. It has been found that both symptomatic & asymptomatic individuals have *Blastocystis* spp. (CDC, 2024).

The prevalence of *Blastocystis* is very high in populations. Based on current estimates, *Blastocystis* spp. infection is considered one of the most common enteric parasitic infections worldwide, potentially affecting over one billion individuals globally (Stensvold and Clark, 2020). A recent retrospective study conducted from 2019 to 2023 in Jeddah-Saudi Arabia, with 7,673 participants, showed that the most predominant parasite in the intestine is *Blastocystis* spp. (48.11%) (Abdalal et al., 2024). A study conducted in Taif, Saudi Arabia, screened 328 children with cancer for intestinal protozoan infections. The results showed that *Blastocystis* spp. was the most frequently detected intestinal parasite among these patients (Hawash et al., 2022). Alqarni et al. (2022) studied intestinal parasite prevalence and detection methods, finding that 52.7% (59/112) of participants were infected. *Blastocystis* spp. was the most prevalent parasite in 86.4% of the positive cases.

Diagnosis of *Blastocystis* spp. in clinical settings depends on detecting *Blastocystis* spp. (usually in the vacuolar form) microscopically in stool samples. Stained slides are preferred more than unstained wet-mount methods to avoid confusion with fecal debris (CDC, 2024). Microscopy has many limitations, including low sensitivity, sample preparation, and the observer’s expertise (Estes, 2016), time consumption, and low specificity due to multi-indistinguishable morphological forms of *Blastocystis* spp. (Padukone et al., 2018). Stool culture, the gold standard method, is more sensitive than microscopy, but it is not used routinely as it is time-consuming. Serodiagnosis has also been developed, but it is not used for routine diagnosis. Molecular methods, PCR, nested-PCR, and RT-PCR are available. Although their efficiency has been demonstrated, implementation requires expert staff, a long operating time, and expensive equipment. Moreover, the accuracy of these assays is a challenge, particularly in samples containing low *Blastocystis* spp. load. For these reasons, it is not ideal for point-of-care (POC) diagnostic purposes (Mahmoud et al., 2023; Wang et al., 2021). Moreover, whilst PCR amplification, sequencing is needed to identify *Blastocystis* spp. subtypes (Padukone et al., 2018). Therefore, an accurate and fast point-of-care testing (PoCT) strategy is essential for the non-invasive diagnosis of *Blastocystis* spp., which is not available.

Loop-mediated isothermal amplification (LAMP) is a molecular technique that has been developed by Notomi et al., (2000). It amplifies DNA with high specificity, efficiency, and rapidity under isothermal conditions. This method employs a DNA polymerase and a set of four to six specially designed primers that recognize distinct sequences on the target DNA. LAMP has several advantages, such as its simplicity in performance, since all it needs is a simple water bath or heating block that provides an isothermal condition. This eliminates the need for advanced instruments in other amplification techniques, such as PCR or RT-PCR (Alhamid et al., 2022). However, neither the microscopy nor the nested PCR (in addition to PCR and RT-PCR) is a perfect standard method for detecting some pathogens, including *Plasmodium falciparum* (Zhang et al., 2022). The simplicity of LAMP is also evident in its method of result analysis. Dyes used in LAMP reactions enable direct visual evaluation of results via naked-eye observation of colorimetric changes or fluorescence under UV light. This eliminates the need for the tedious gel electrophoresis procedure required by PCR (Wong et al., 2018).

This study aimed to develop fluorometric and colorimetric LAMP, namely fLAMP and cLAMP, techniques as an alternative PoCT for the diagnosis of *Blastocystis* spp. The assays were tested on 194 symptomatic and asymptomatic individuals and were compared with standard RT-PCR protocols. Comparative analyses, including sensitivity, specificity, and accuracy, were performed. This is the first study describing colorimetric and fluorometric LAMP-based assays in the diagnosis of *Blastocystis* spp.

## MATERIAL AND METHODS

### Sample collection and copro-DNA extraction

The stool samples (n = 197) were collected from patients who were admitted to the King Fahd University Hospital (KFUH) and the Family and Community Medicine Center (FCMC) of Imam Abdulrahman bin Faisal University (IAU) in Dammam, Saudi Arabia, between August and October 2024. The patients had gastrointestinal complaints, such as diarrhea, vomiting, abdominal pain, constipation, and epigastric pain. The Institutional Review Board (IRB) at IAU approved the study with an IRB number of IRB-2024-13-530.

After collecting the samples, copro-DNA extraction was performed in a BSL-2 cabinet using the QIAamp DNA Stool Mini Kit (Qiagen, Germany). The quality and quantity of the extracted DNA were assessed by spectroscopic measurement of the samples’ absorbance at 260 nm and 280 nm, and by determining the 260/280 ratio using a Nano Drop 2000 (Thermo Scientific).

### Alignment of *Blastocystis* spp. genome sequences and primer design

Seven of the most common subtypes of *Blastocystis* spp.—namely ST1 to ST7—were selected from different hosts, including human, rat, pig, turkey, and chicken. The sequence information for these subtypes was derived from Santín et al. (2011). At least three sequences belonging to each subtype were retrieved from the NCBI nucleotide database (https://www.ncbi.nlm.nih.gov/nucleotide/). The sequences were aligned using the Clustal Omega tool (https://www.ebi.ac.uk/jdispatcher/msa/clustalo) and visualized with the Mview tool (EMBL-EBI) (https://www.ebi.ac.uk/jdispatcher/msa/mview?stype=protein). LAMP oligonucleotides were designed using the PrimerExplorer V5 program (https://primerexplorer.jp/e) by targeting the conserved genomic sequences of the *18 SSU rRNA* gene. The positions of each primer (LF, LB, F3, F2, F1, B1c, B2, and B3) showed a 100% consensus for each subtype. The OligoAnalyzer tool from Integrated DNA Technologies (IDT) (https://eu.idtdna.com/pages/tools/oligoanalyzer) was used to verify that no undesirable secondary structures, such as homodimers, heterodimers, or hairpins, would form with the oligonucleotides.

The lyophilized and desalted oligonucleotides were manufactured by Alpha DNA (Montreal, Canada) (http://www.alphadna.com). The final concentrations of the oligonucleotides (10×) in a 25 μL reaction volume were adjusted to be 2 μM for F3 and B3, 8 μM for LF and LB, and 16 μM for FIP and BIP. The oligonucleotide sequences used in this study are displayed in **Table 1**.

**Table 1.**
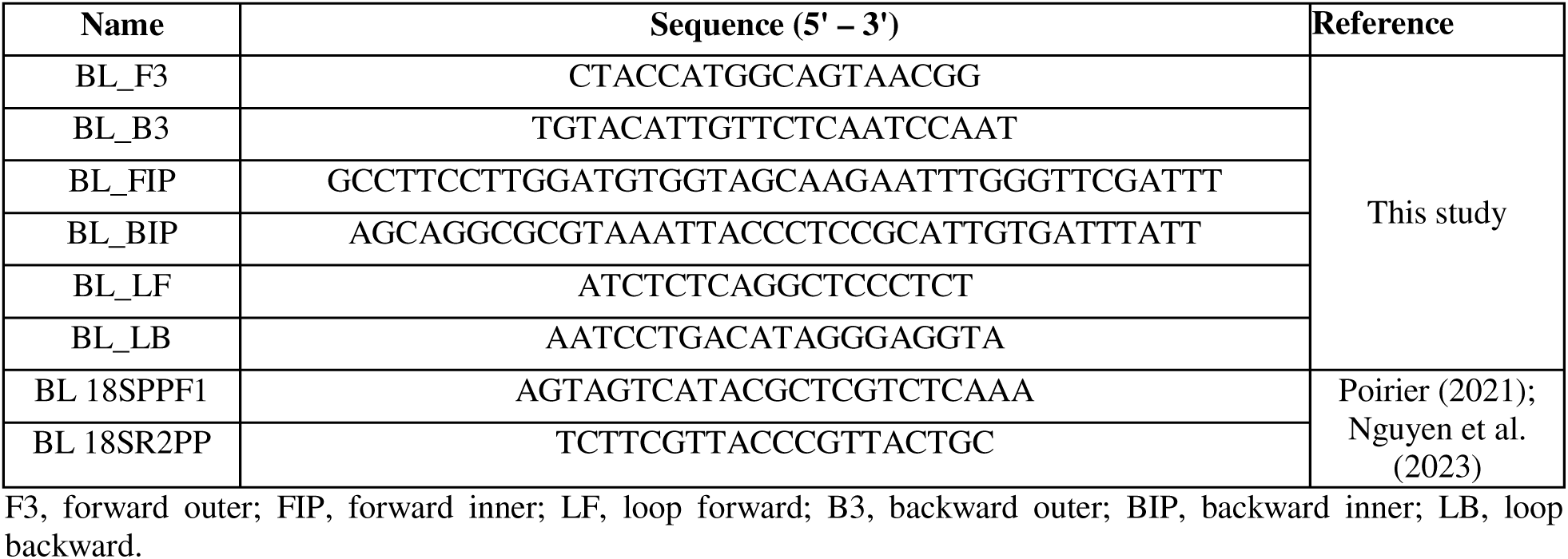
The sequences of primers used in this study.

### Real-time PCR analysis

Real-time PCR (RT-PCR) analysis was conducted to verify the presence of *Blastocystis* spp. in the stool sample. RT-PCR was performed using *Blastocystis*-specific forward and reverse primers, which target the *18 SSU rRNA* gene **(Table 1)**. The RT-PCR was conducted by combining the following components: 5x concentrated SolisFast^®^ SyberGreen Master mix (Solis BioDyne, Estonia), 10 mM of each primer, 1 µL of copro-DNA (or dH□O for a non-template control, NTC), and ddH□O to a final volume of 20 µL. The reactions were performed using an Applied Biosystems 7500 Fast Real-Time PCR (Waltham, MA, USA) machine. The reaction conditions were set as follows: an initial denaturation at 95 °C for 10 minutes, followed by 95 °C for 10 seconds, 60 °C for 20 seconds, and 72 °C for 10 seconds (40 cycles). Following amplification, melting curve analysis was applied with settings of 60-95 °C, with 0.1 °C increments for each reaction. A threshold cycle (Ct or Cq) score above 35 (Ct > 35) was accepted as a negative result. *Blastocystis*-positive specimens were identified if the Ct value was ≤ 35, accompanied by a sigmoidal amplification curve and a single peak around 80-84 °C in the melting curve analysis.

### Fluorometric LAMP reaction

The fluorometric LAMP (fLAMP) reactions were conducted in 0.2 mL reaction tubes with a total volume of 25 µL. 20x Green Fluorescent Dye (Lucigen, USA) was used as the fluorometric reagent. The master mix contained 10x isothermal buffer (New England BioLabs, NEB), 100 mM MgSO□ (NEB), 10 mM deoxynucleotide (dNTP), Green Fluorescent Dye (20x), *Bst* 2.0 polymerase (8,000 U/µL) enzyme, fLAMP primers mixture (10x), template DNA (10-50 ng), and ddH□O to a final volume of 25 µL. ddH□O was used for the negative control (NC) or non-template control (NTC) reactions. To improve sensitivity, Betaine, which is known to improve the DNA-primer interaction, was added to the reaction mixture in varying concentrations (0, 0.4, and 0.8 M). The fLAMP reactions were performed in a thermocycler (Applied Biosystems 7500 Fast Real-Time PCR, ThermoFisher Scientific, Waltham, MA, USA), with the reaction conditions set to 65 °C for 80 cycles (each cycle lasting 45 seconds).

### Colorimetric LAMP reaction

The colorimetric LAMP (cLAMP) reaction mixture consists of the following components: 2x WarmStart® cLAMP master mix with UDG (NEB), cLAMP primer mix (10x), template DNA (10-50 ng), and ddH□O to a final volume of 20 µL. The mixture was placed in a thermocycler (Techne TC-512, Germany), and the reaction was initiated by setting the conditions at 65 °C for 55 minutes. The reactions were terminated by removing the tubes from the thermocycler and storing them at -20 °C. The color change was monitored at 5-minute intervals.

### Evaluation of assay sensitivity and accuracy

To assess the reliability of the fluorometric and colorimetric assays, their diagnostic performance was evaluated against RT-PCR. For this study, RT-PCR was designated as the definitive reference standard for classifying all samples as either positive or negative. A confusion matrix was constructed for each assay to quantify the number of true positives (TP), true negatives (TN), false positives (FP), and false negatives (FN) results. The diagnostic metrics of sensitivity, specificity, accuracy, positive predictive value (PPV), and negative predictive value (NPV) were subsequently calculated using the data from the confusion matrices. The formulas and methodology for these calculations were adapted from established literature (Alhamid et al., 2023). This comprehensive approach ensures a rigorous and robust evaluation of the diagnostic capabilities of each assay.

### Validation of LAMP Products

The integrity of both cLAMP and fLAMP amplification products was validated using agarose gel electrophoresis. A 2% agarose gel, prepared with a DNA stain like VisualaNA (Molequle-On, New Zealand), was used to separate the DNA fragments. The gel was run in an electrophoresis system at 100 V for 45 minutes. Following electrophoresis, the gel was visualized using a UV transilluminator (ChemiDoc™ XRS+ System with Image Lab™ Software, Bio-Rad, USA). Successful amplification in positive samples was confirmed by the presence of a characteristic ladder-like pattern of DNA bands. This unique pattern is a hallmark of LAMP amplification, resulting from the self-priming and strand-displacing activity of the polymerase, which creates a series of concatemers of various sizes.

In addition to gel electrophoresis, the fLAMP reactions were also monitored using fluorescent dye. To each reaction tube, 1 µL of a 20x Green Fluorescent Dye (Lucigen) was added. The tubes were then observed under UV light. Positive reactions were identified by a noticeable fluorescence intensity, indicating the presence of a high concentration of amplified DNA. Negative reactions remained non-fluorescent or showed minimal background fluorescence. This method offers a rapid and convenient way to visually confirm the presence of amplification under UV light.

### Cloning of the *18 SSU rRNA* gene into a plasmid vector

To avoid co-infection and evaluate the sensitivity of LAMP primers, *18 SSU rRNA* gene was cloned into a plasmid vector. A 490 bp segment of the *18 SSU rRNA* gene was integrated into pET21/pGSI plasmids. The cloning utilized NotI and EcoRI restriction enzymes (New England BioLabs, UK) for digestion before ligation. The ligated plasmid (50 µL) was then introduced into competent *E. coli* JM109 via heat shock (42□C for 45 seconds). After recovery in SOC medium, cells were spread (50 µL per plate) on agar plates containing ampicillin (50 µg/mL) and incubated overnight at 37□C. Positive colonies were chosen and validated through PCR. The plasmid was subsequently purified using the QIAprep Spin Miniprep Kit (Qiagen, Germany), quantified with a NanoDrop 2000 (Thermo Sci), and reserved at -20□C for its use as a DNA template in determining the limit of detection (LoD).

### Calculation of limit-of-detection (LoD)

The LoD was determined using serial dilutions of plasmid DNA containing a part of *18 SSU rRNA* gene (490 bp) and copro-DNA with a pre-determined concentration using NanoDrop 2000c (Thermo Sci.). At least three replicates were used. The fLAMP and cLAMP reactions were carried out in a 7500 Fast Real-Time PCR System (Thermo Sci., MA, USA). The reaction conditions were set to 65 °C for 80 cycles (45 seconds for each cycle). Subsequently, the samples were loaded onto a 2% agarose gel for validation of positive or negative amplifications and subjected to electrophoresis in an Analytik Jena unit at 100 V for 45 minutes. Finally, the gel was monitored under a UV-trans illuminator (Bio-Rad, USA).

### Specificity assay

To test the LAMP primer’s specificity, we utilized the most common parasites found in the stool samples, including *Cryptosporidium* (Pedraza-Díaz et al., 2001; Spano et al., 1997), *Giardia* (Read et al., 2002), *Helicobacter pylori* (Tombuloglu, 2025), *Entamoeba* (Foo et al., 2012), *Dientamoeba* (Stark et al., 2005). The copro-DNA samples were validated for the positivity of these species by using nested-PCR, according to the suggested protocols. Then *Blastocystis* spp. RT-PCR and LAMP primers were tested against the same samples to confirm their negativity with *Blastocystis* spp.

### Statistical Analysis

To evaluate the statistical diagnostic performance of the LAMP assays, the results obtained from cLAMP (n = 151) and fLAMP (n = 194) tests were independently compared against the corresponding results from the reference standard RT-PCR. This comparative analysis enabled the calculation of diagnostic sensitivity, specificity, accuracy, PPV, and NPV for each LAMP assay.

## RESULTS

### Determination of *Blastocystis* spp. in clinical samples

Copro-DNA concentrations range from 4.4 to 593.4 ng/µL. DNA purity was assessed through the A_260_/A_280_ ratio, and the samples within the range of 1.70 to 2.00 were accepted as standard quality, while those out of this range were excluded from further studies. The total number of samples that correspond to these criteria was 197. Therefore, the following experiments were conducted with this sample size.

Before the application of the LAMP assay, the presence/absence of target *Blastocystis* DNA in the clinical samples (n = 197) was verified by using RT-PCR and melting curve analysis using *Blastocystis*-specific primers (**Table 1**). The RT-PCR reactions produced a sigmoidal amplification curve (Ct ≤ 35), and a single melting point peak (between 80-84 °C) was noted as *Blastocystis* spp. positive. On the other hand, the negative samples and no-template control (NTC) produced a parallel line (**Figure 1A**). Accordingly, RT-PCR yielded 102 *Blastocystis* spp. positive and 92 *Blastocystis* spp. negative results. In addition, three samples were noted as inconclusive, since they did not produce a definitive result. Thus, they were excluded from the final sample set.

**Figure 1.**
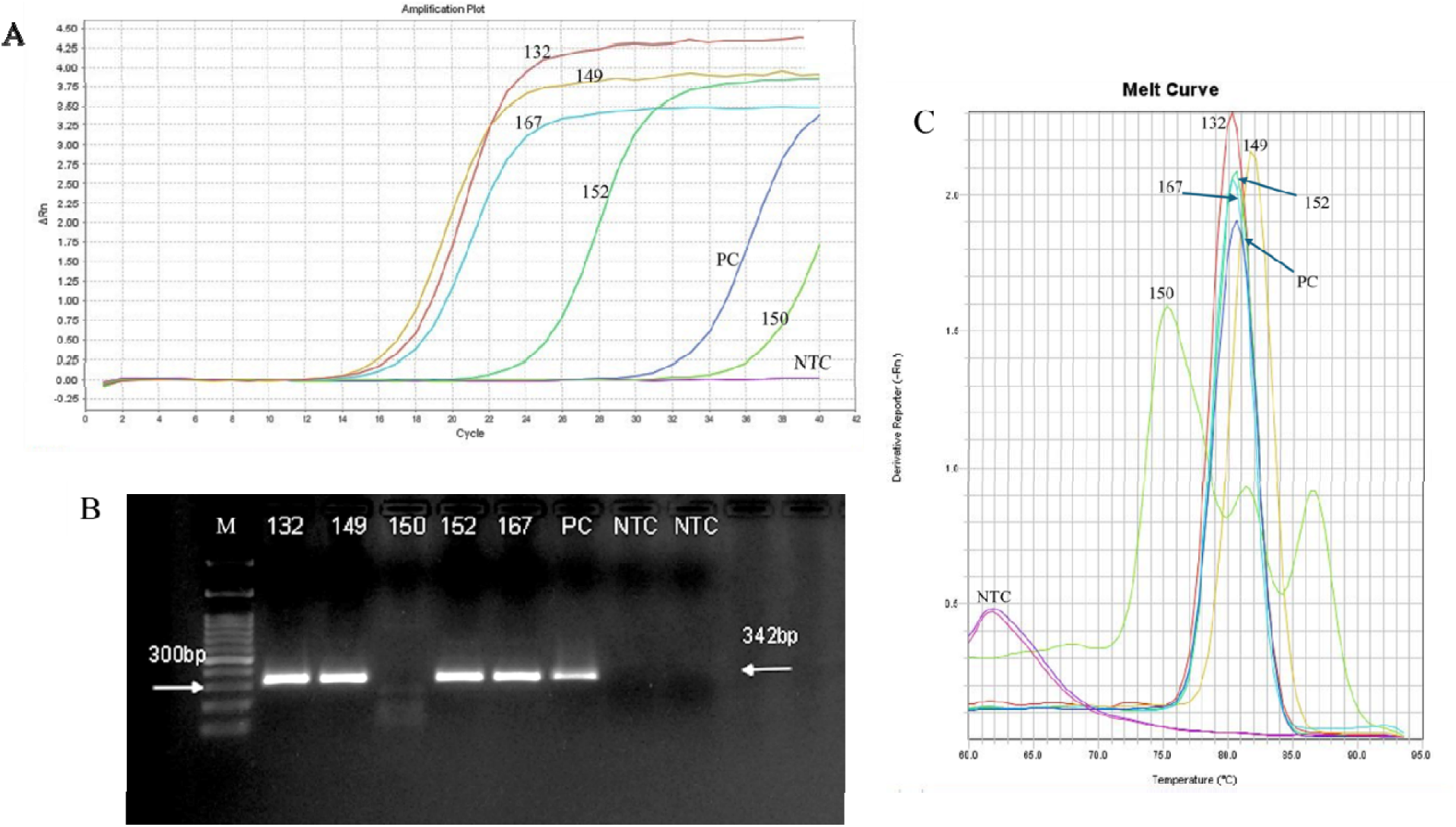
Representative figures show the real-time PCR analysis of the *Blastocystis* spp. *18 SSU rRNA* gene. **(A)** Real-time PCR amplification plot. Amplification plots for positive sample (132, 149, 152, 167, and PC) exhibit Ct values ≤ 35, while the negative control (NTC) and negative sample (150) showed no amplification or late amplification (Ct > 35). **(B)** Agarose gel electrophoresis (2%) result after loading RT-PCR products. Positive samples and PC show a clear and single targeted band size (342 bp), while negative samples and NTC show no bands. **(C)** Melting curve analysis of the same samples. Positive samples showed melting curves within a temperature range from 80 to 84 as a single peak. (M = 100 bp size marker).

The amplified RT-PCR gene products were also visualized in 2% agarose gel, confirming a single band (∼342 bp) in positive samples, while no bands were observed in NTC and negative results (**Figure 1B**). Since the length of the target gene amplicon may vary in base pairs due to the nucleotide variance in different subtypes, the melting point of each amplicon could be varied (**Figure 1C)**.

### Detection of *Blastocystis* spp. by fluorometric LAMP assay

**Figure 2A** and **Figure S1** depict exemplified fLAMP reactions using an RT-PCR system equipped for fluorescence detection. Successful amplifications of the target *18 SSU rRNA* gene were denoted by the characteristic sigmoidal curve observed in positive samples. These samples exhibited Ct values ranging from 20 to 53, suggesting a high initial concentration of the target sequence and a fast amplification of target DNA. Conversely, in samples where the target DNA was absent (including the negative control), the fLAMP analysis demonstrated no significant increase in fluorescence signal throughout the reaction. Furthermore, these samples did not exhibit any aberrant amplification products, confirming the specificity of the assay for the intended target sequence.

**Figure 2.**
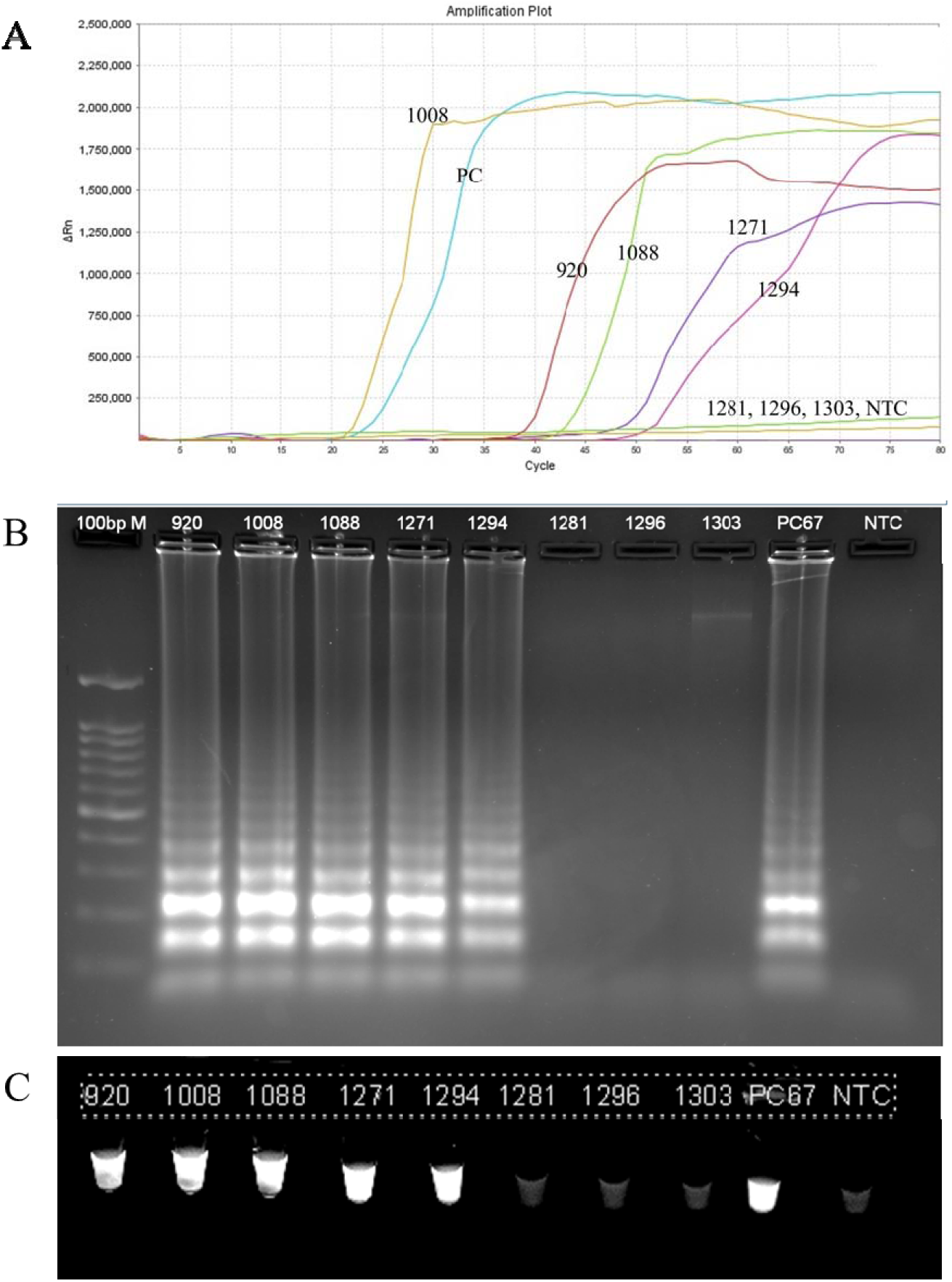
Isothermal amplification of the target *18S SSU rRNA* gene by using fluorometric LAMP. **(A)** Fluorometric LAMP amplification plot. Positive samples (920, 1008, 1088, 1271, 1294, and PC67) showed obvious sigmoidal amplification curves, while negative samples (1281, 1296, 1303, and NTC) produced no amplification curve. **(B)** Agarose gel electrophoresis (2%) after loading fluorometric LAMP products of the same samples. Positive samples showed a ladder-type banding pattern confirming successful amplification, while negative samples showed no banding pattern. **(C)** Observation of fLAMP products under UV transilluminator. (M = 100 bp size marker).

**Figure 2B** displays the gel electrophoresis results of the fLAMP products. As illustrated in the figure, a ladder-like banding pattern was evident for the positive reactions. In contrast, the negative tubes did not produce this banding pattern, corroborating the fluorometric amplification curve results. The LAMP amplicons were also visualized under a UV transilluminator (**Figure 2C; Figure S1**). The result is in line with fLAMP and agarose gel electrophoresis.

### Detection of *Blastocystis* spp. by colorimetric LAMP assay

Colorimetric LAMP (cLAMP) reaction was performed in a thermocycler set at 65 °C with lid temperature of 100 °C. The reactions were observed every five minutes. To avoid misamplification and false positive results, the reaction was stopped after 55 minutes. **Figure 3** and **Figure S2** depict the color development in *Blastocystis* spp. samples, which exemplified test groups, include 29 and 30 samples, respectively, with PC and NC reaction tubes. The initial reaction color at 0 min was red, which turned yellow in *Blastocystis*-positive reaction tubes, and remained red in negative ones. As expected, the PC and NC tubes appeared yellow and red, respectively, without misamplification. These results show the successful detection of *Blastocystis* spp. in positive samples by using the cLAMP method.

**Figure 3.**
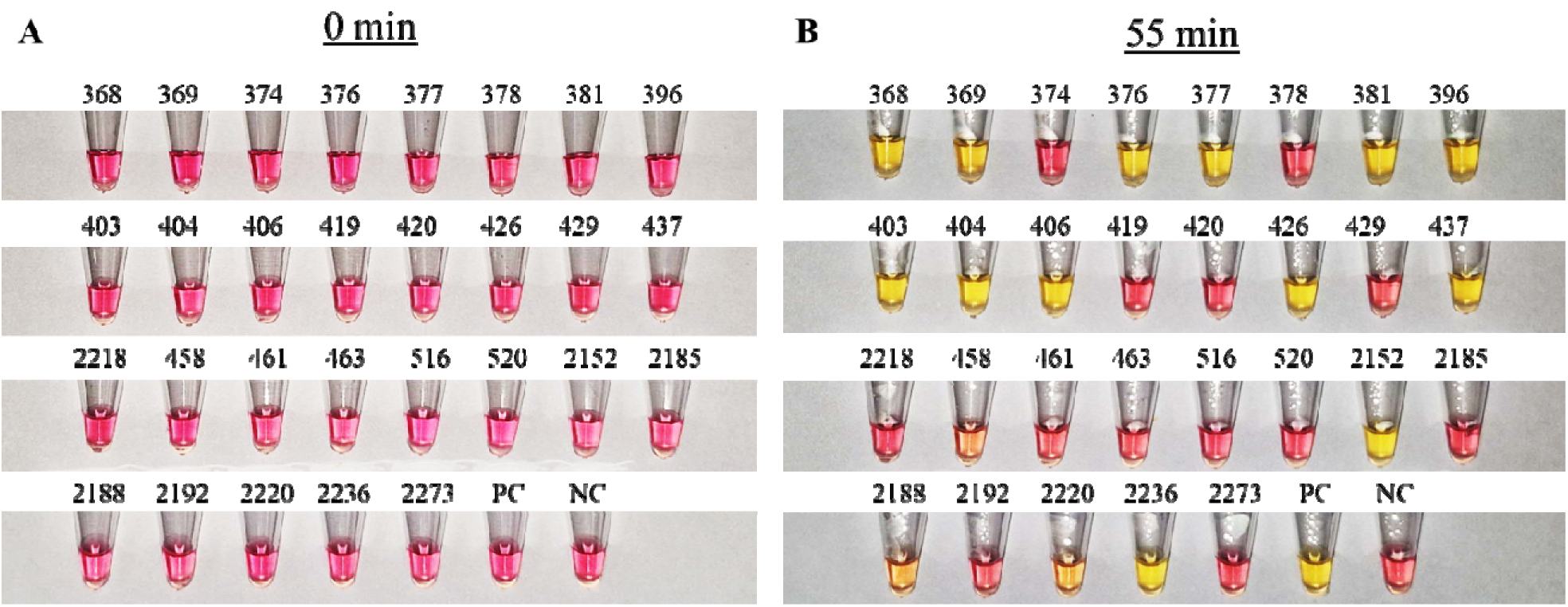
Colorimetric detection of *Blastocystis* spp. Comparison of the reactions **(A)** before (0 minutes) and **(B)** after the reaction (55 minutes). This group includes 29 unknown samples with positive control (PC) and negative control (NC) reaction tubes.

### Detection kinetics of colorimetric and fluorometric LAMP assays

The color development in the cLAMP reaction over time is shown in **Figure 4**. The figure shows six randomly selected *Blastocystis* spp. positive and negative samples that have been verified by RT-PCR and fLAMP assays. The color development was monitored every five minutes starting after 20 to 55 minutes. It was obvious from the results that the color development for some *Blastocystis* spp. positive samples started within 20 minutes, which can be distinguished between the positive and negative tubes. And the red to yellow color change was fully apparent after 30 minutes. The reaction was monitored for 55 minutes. However, the NC with the other negative sample tubes remained red, as expected. These results show the specificity and sensitivity of the cLAMP assay in diagnosing *Blastocystis* spp.

**Figure 4.**
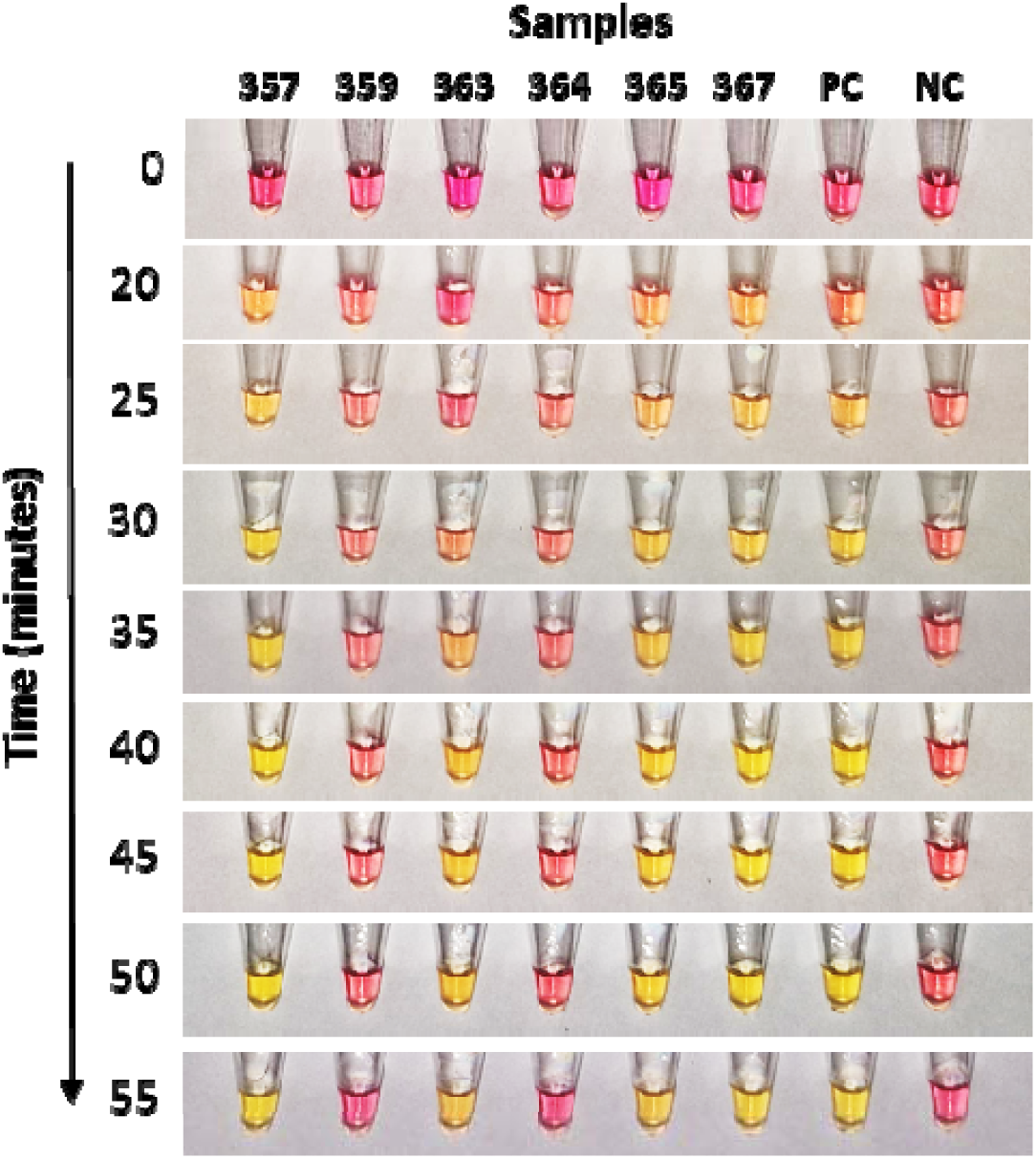
The color development in the colorimetric LAMP reaction over time. The figure shows six randomly selected *Blastocystis*-positive or negative samples, in addition to a positive control (PC) and a negative control (NC). The color development was monitored every five minutes starting from 20 minutes to 55 minutes.

The detection time for the positive samples in the fLAMP reaction ranged from 20 to 50 minutes, with an average detection time of 28.2 minutes. Compared to the RT-PCR and fLAMP results over 194 samples, 92.27% of the results were consistent. The inconsistency between the two assays (7.73%) could be attributed to various factors that might affect the reactions, such as DNA quality and concentration, or the presence of any PCR or LAMP inhibitors (further discussed in the discussion section).

### Comparison of the time-to-detection threshold

The average time-to-detection (TT) values for the cLAMP (n = 151), fLAMP (n = 194), and RT-PCR (n = 194) assays were computed **(Figure 5)**. The cLAMP assay had a mean TT of 29.2 minutes, which is significantly faster than the reference RT-PCR assay’s mean TT of 36.2 minutes. The fLAMP assay was even faster, with an average TT of just 24.0 minutes. It’s important to note that the TT for both RT-PCR and fLAMP is based on the threshold cycle (Ct) value, which represents the point where the amplification curve crosses the detection threshold. While the TT for RT-PCR was 36.2 minutes, the entire one-and-a-half-hour protocol, including melting curve analysis, should be considered when comparing overall assay completion times. Overall, these results show that both the fLAMP and cLAMP assays provide a reduced time to detection of *Blastocystis* spp. compared to conventional RT-PCR.

**Figure 5.**
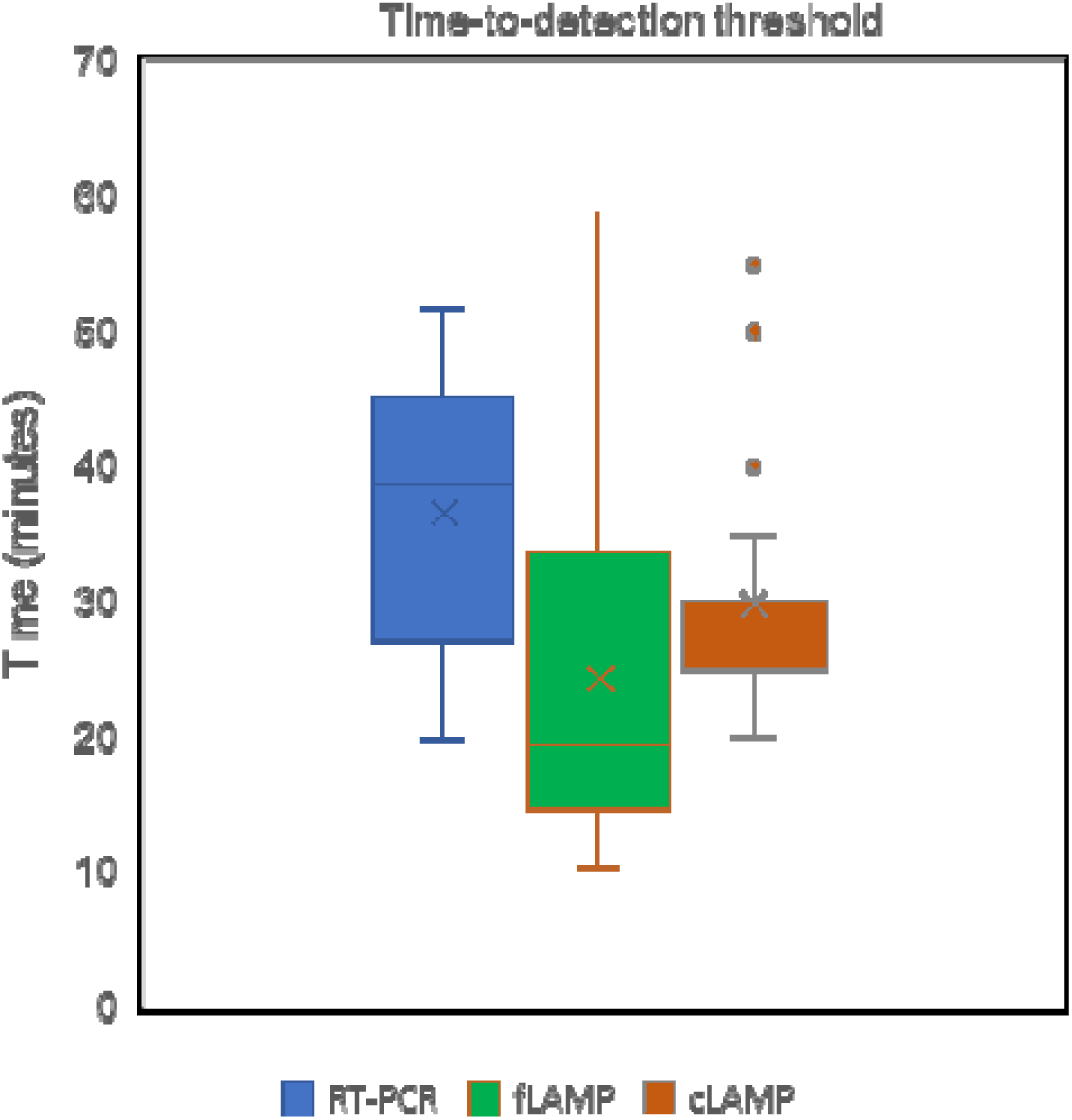
Comparative analysis of the mean time-to-detection threshold (TT) of the three assays: RT-PCR, fLAMP, and cLAMP. The dashed line indicates the median of each assay.

### Sensitivity and specificity of fluorometric and colorimetric LAMP assays

A cohort of 194 clinical specimens underwent comparative testing using both the reference RT-PCR and the fLAMP assays. **Table 2** shows the number of true positives (TP), true negative (TN), false positives (FP), and false negatives (FN) results compared to the RT-PCR. Of the 194 samples, 179 fLAMP results agreed with RT-PCR, with six FP and nine FN. The results revealed that the fLAMP assay has 91.43% sensitivity, 93.26% specificity, and 92.27% accuracy. The PPV was calculated to be 94.12%, while the NPV was 90.22% (**Table 2)**.

**Table 2.** Diagnostic yield of fLAMP and cLAMP compared to standard RT-PCR.

|  |  | fLAMP |  |  | cLAMP |  |  |
| --- | --- | --- | --- | --- | --- | --- | --- |
|  |  | (+) | (-) | Total | (+) | (-) | Total |
| RT-PCR | (+) | 96 (TP) | 6 (FN) | 102 | 68 (TP) | 3 (FN) | 71 |
|  | (-) | 9 (FP) | 83 (TN) | 92 | 12 (FP) | 68 (TN) | 80 |
|  | Total | 105 | 89 | 194 | 80 | 71 | 151 |

\* true positive (TP), true negative (TN), false positive (FP), and false negative (FN).

In addition, a total of 151 clinical specimens were subjected to parallel testing using both the reference standard RT-PCR and cLAMP assays (**Table 2)**. Of the 151 samples, 136 cLAMP results agreed with the reference RT-PCR method, with 12 FP and three FN. The comparison revealed that the cLAMP assay has 95.77% sensitivity, 85.00% specificity, and 90.07% accuracy. The PPV was calculated to be 85.00%, while the NPV was 95.77%.

### Determination of limit-of-detection

To calculate the limit of detection (LoD), two different positive control DNA samples with known initial DNA concentrations were used. The first is formed by pooling DNA samples from five different pre-determined *Blastocystis* spp. DNA specimens derived from the clinical specimens (tested with RT-PCR, fLAMP, and cLAMP), and their concentration was spectrophotometrically determined. It should be noted that this calculation determines the copro-DNA concentration in the sample that includes *Blastocystis* spp. together with other possible DNA samples belonging to different organisms.

The initial copro-DNA concentration was 14.3 ng/µL, and it was serially diluted up to 10^6^ times (14.3, 14.3×10^-1^, 14.3×10^-2^, 14.3×10^-3^, 14.3×10^-4^, 14.3×10^-5,^ and 14.3×10^-6^). The DNA samples were then subsequently analyzed via fLAMP and cLAMP and verified with gel electrophoresis and fluorescent determination under UV light (**Figures 6 and 7**). The experiments were performed in triplicate. For the copro-DNA, the LoD of the fLAMP assay was determined as 14.3×10^-4^ ng/µL, as this was the lowest concentration at which consistent amplification was observed across all replicates (**Figure 6)**. This corresponds to 1,430 fg/µL of copro-DNA. The amplicons were also monitored by loading them onto 2% agarose gel and under UV light, as shown in **Figure 6B** and **C**, respectively. On the other hand, LoD of the cLAMP assay was 1.43×10^-2^, which was relatively lower compared to the fLAMP assay **(Figure 7)**. This could be related to the color development, which might have been affected by possible LAMP-interfering substances, such as salts, copro-DNA, etc.

**Figure 6.**
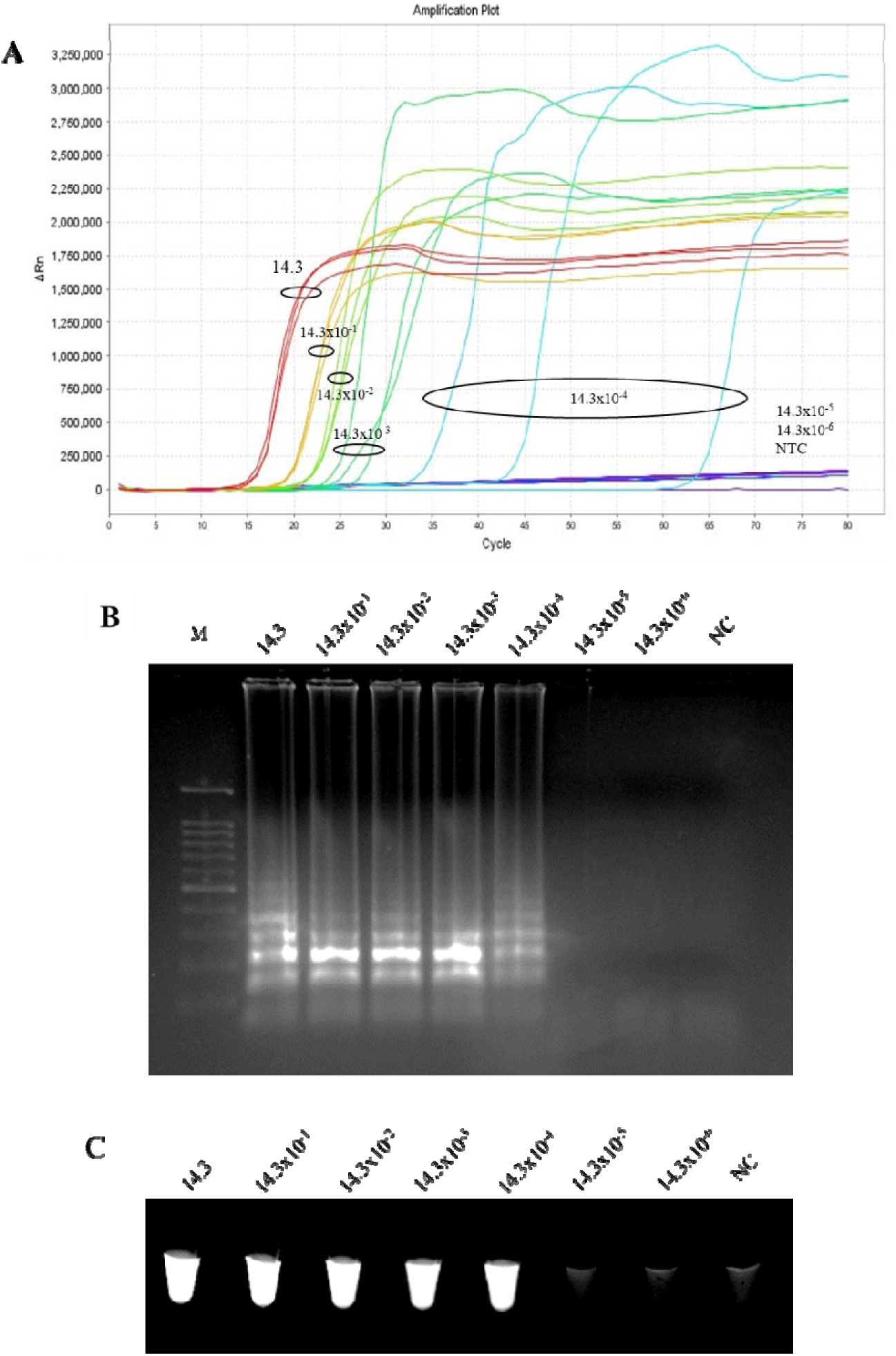
Determination of the sensitivity of the fluorometric LAMP assay. **(A)** Amplification plot shows serially diluted copro-DNA samples (14.3, 14.3×10^-1^, 14.3×10^-2^, 14.3×10^-3^, 14.3×10^-4^, 14.3×10^-5,^ and 14.3×10^-6^ ng/µL). Concentrations of 14.3×10^-5^, 14.3×10^-6,^ and NTC gave no amplification plots. **(B)** Agarose gel electrophoresis (2%) after loading fluorometric LAMP products. Positive samples showed a ladder-type banding pattern, confirming successful amplification. **(C)** fLAMP tubes under UV transilluminator. (M = 100 bp size marker).

**Figure 7.**
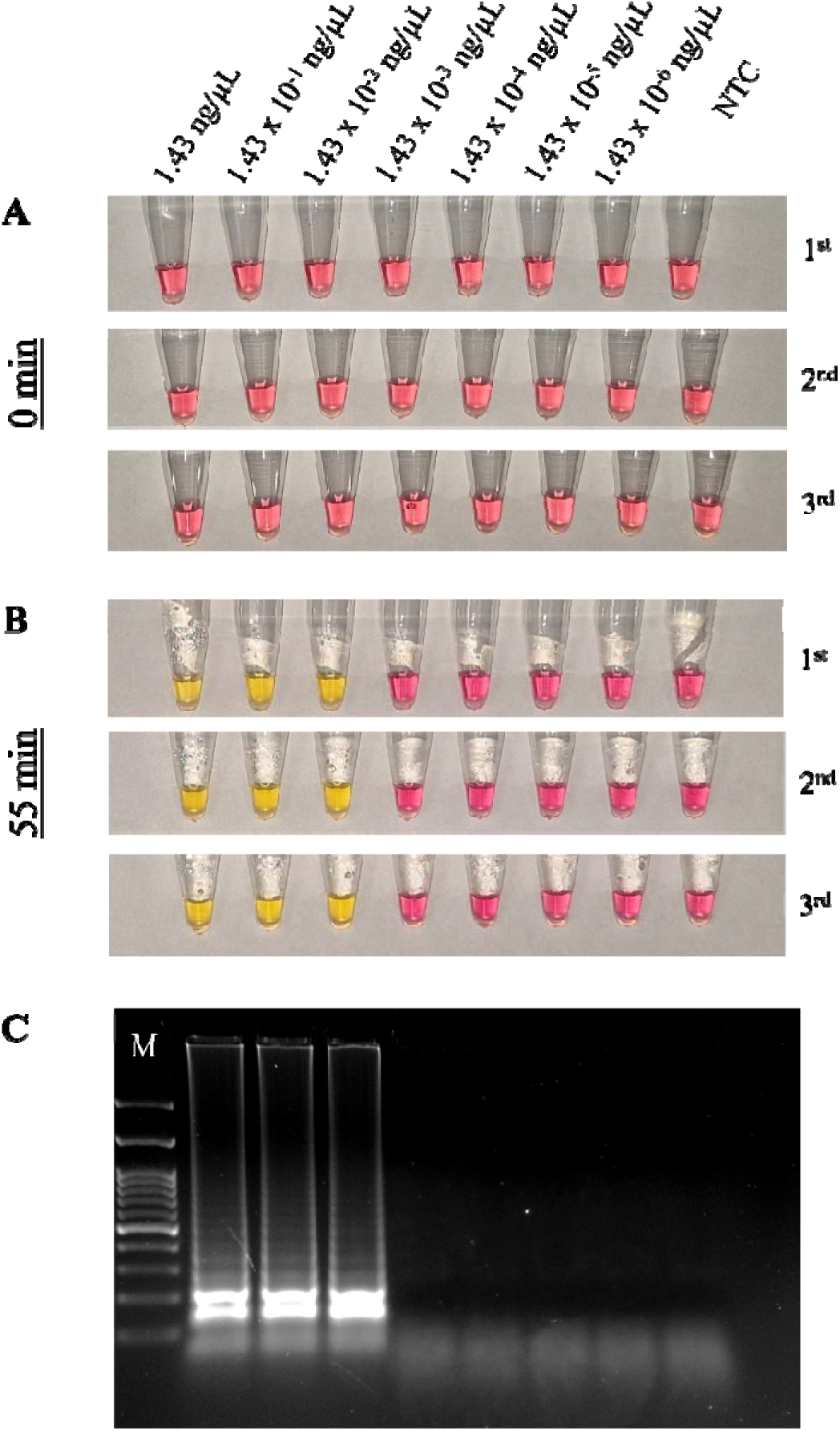
Determination of the sensitivity of the colorimetric LAMP assay. The color development in serially diluted copro-DNA samples (1.43, 1.43×10^-1^, 1.43×10^-2^, 1.43×10^-3^, 1.43×10^-4^, 1.43×10^-5^, and 1.43×10^-6^ ng/µL) **(A)** before (0 min) and **(A-B)** after (55 min) reaction. **(C)** Agarose gel electrophoresis (2%) after loading colorimetric LAMP products. Positive samples showed LAMP ladder-type banding pattern, confirming successful amplification. (M = 100 bp size marker).

In addition, the LoD of the fLAMP and cLAMP assays was computed by using a plasmid vector containing a piece of the *18 SSU rRNA* gene. For this purpose, 490 bp of the *SSU rRNA* gene was synthesized and cloned into the pUC21/pGSI plasmid, and its concentration wa determined. This sample provides real *Blastocystis* spp. nucleic acid, avoiding other organismal DNA samples residing in the copro-DNA samples.

The initial pDNA concentration was 5 ng/µL, and it was serially diluted up to 10^6^ time (5, 5×10^-1^, 5×10^-2^, 5×10^-3^, 5×10^-4^, 5×10^-5^, and 5×10^-6^). The DNA samples were then subsequently analyzed via fLAMP and cLAMP with three replicates. For the pDNA, the LoD of the fLAMP and cLAMP assays were determined to be 5×10^−6^ ng/µL, corresponding to 5 fg/µL (**Figures 8 and 9**). This indicates that the assay is sensitive at the femtogram level. The chart in **Figure 8C** shows the consistency of triplicate experiments and amplification efficacy of fLAMP reactions with an R^2^ value of 0.9969.

**Figure 8.**
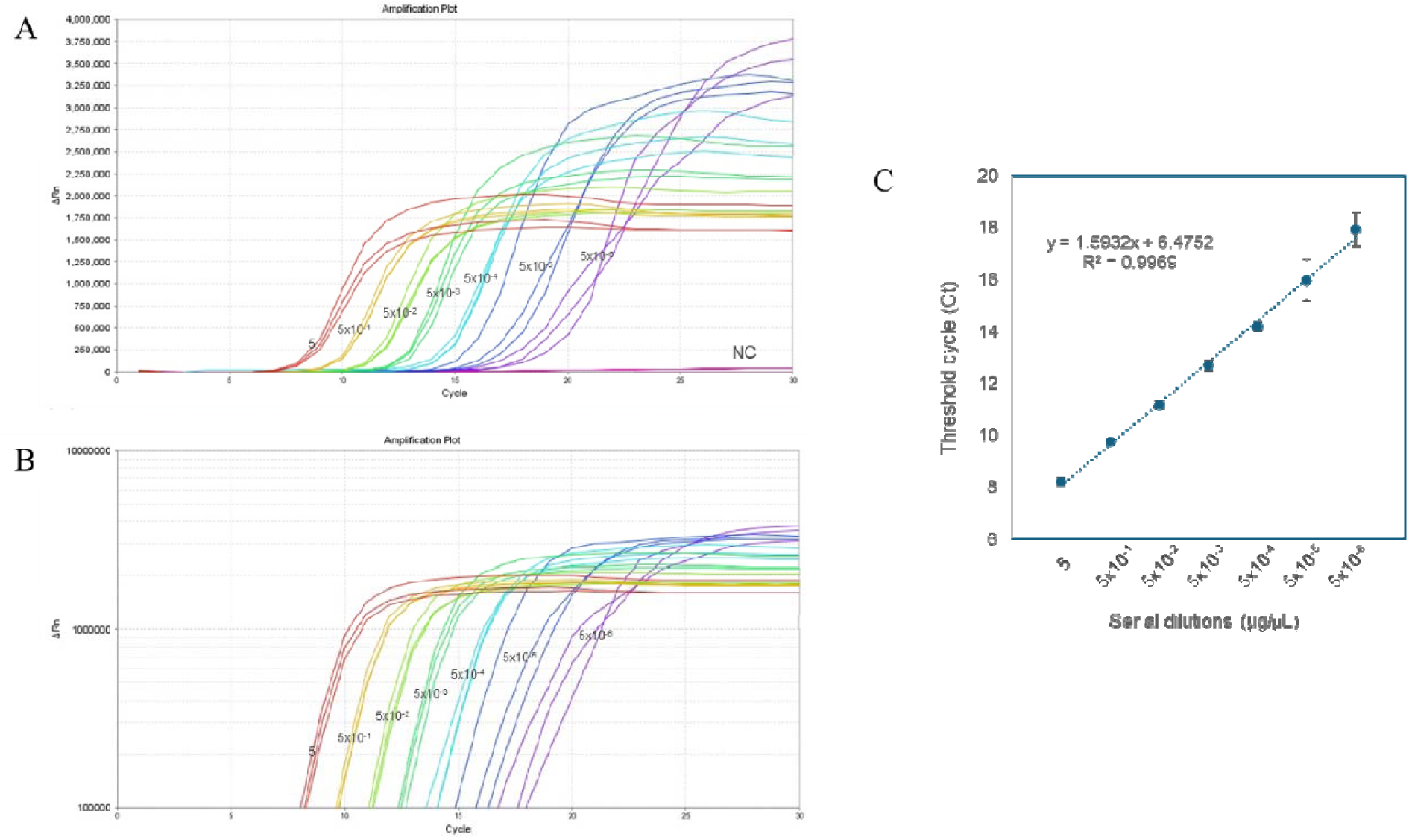
Fluorometric LAMP results for serially diluted plasmid DNA containing the target *18 SSU rRNA* gene (490 bp). **(A)** fLAMP amplification plot showing samples with concentrations; 5, 5×10^-1^, 5×10^-2^, 5×10^-3^, 5×10^-4^, 5×10^-5^, and 5×10^-6^ ng/µL. The negative control (NC) gave no amplification plots. **(B)** The logarithmic representation of the amplification curves with three technical replicates. **(C)** The consistency of triplicate experiments and amplification efficacy of fLAMP reactions. Error bars represent the standard deviation over three technical replicates.

**Figure 9.**
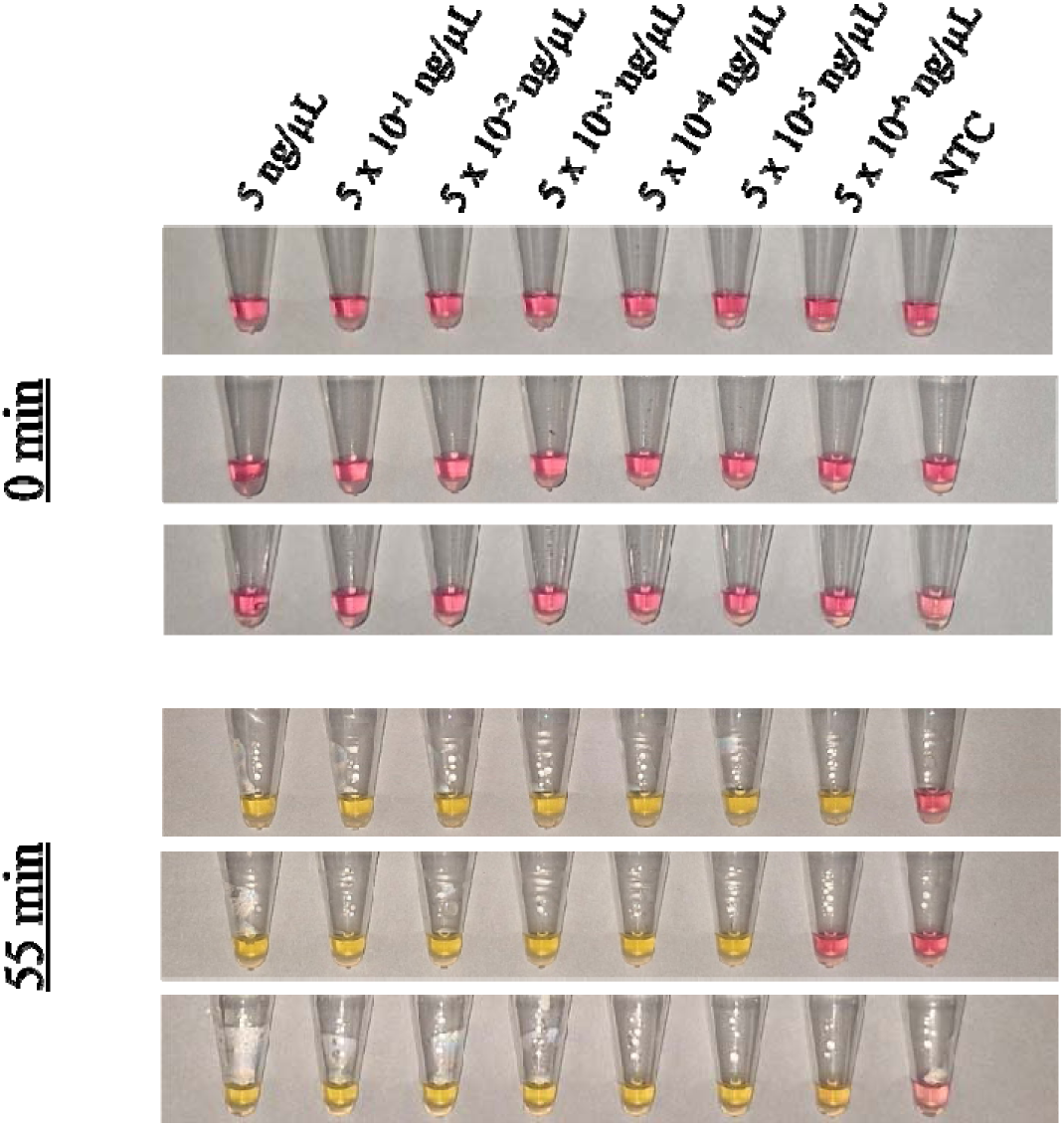
Determination of cLAMP assay analytical sensitivity for serially diluted plasmid DNA containing the target *18 SSU rRNA* gene (490 bp). The color detection shows serially diluted plasmid DNA with concentrations 5, 5×10^-1^, 5×10^-2^, 5×10^-3^, 5×10^-4^, 5×10^-5,^ and 5×10^-6^ ng/µL. All dilutions show a positive color change up to 5 femtograms.

### LAMP Primers Specificity

The specificity test showed that the designed *Blastocystis* LAMP primers have no cross-reactivity with *Cryptosporidium*, *Giardia*, *Entamoeba*, *Dientamoeba*, *H. pylori,* and human DNA (**Table S1)**. So, other parasites’ DNA did not amplify using the LAMP reaction.

## DISCUSSIONS

Significant research efforts are directed towards developing alternative molecular and serological methodologies to achieve cost-effective and timely detection of *Blastocystis* spp. The challenges associated with detecting *Blastocystis* spp. in fecal samples have led to uncertainty and misunderstandings regarding its life cycle, host specificity, and pathogenicity (Santín et al., 2011). On top of it, neither microscopy nor nested-PCR (in addition to PCR and RT-PCR) is a perfect gold standard method in the diagnosis of *Blastocystis* spp.

The loop-mediated isothermal amplification (LAMP) reaction offers several advantages for point-of-care (POC) testing. It proceeds under isothermal conditions within a single reaction vessel, eliminating the need for complex thermocycling instruments. Furthermore, LAMP assays employ straightforward detection methods, are cost-effective, and provide rapid results, making them well-suited for decentralized diagnostic applications (Alhamid et al., 2022; Notomi et al., 2000; 2015). Moreover, accurate, rapid, and accessible diagnostic assays are paramount for controlling the propagation of *Blastocystis*-related conditions. While RT-PCR remains a reference standard assay, significant research efforts are directed towards developing alternative molecular and serological methodologies to achieve cost-effective and timely detection of *Blastocystis* spp.

Rivera & Ong (2013) demonstrated the feasibility of LAMP for detecting *Entamoeba histolytica*, noting its high specificity and visual readout. They further confirmed that the LAMP technique is highly specific to its target, showing no cross-reactivity with other gastrointestinal parasites, which reinforces its utility as a rapid diagnostic tool. In recent research, Peng et al., (2024) achieved a rapid and highly sensitive cLAMP assay for Mpox virus using pseudovirus clones and an integrated device, visually confirming the assay results with the naked eye. Similarly, Prathyusha et al., (2025) revealed a novel cLAMP assay for the rapid, field-level detection of *Theileria orientalis*. Their work demonstrated the effectiveness and specificity of the cLAMP assay, highlighting its suitability for field-level diagnosis in resource-limited settings. The use of visual dyes and simplified protocols in these studies aligns with our goal of developing accessible molecular diagnostics. Finally, a study by Robert-Gangneux et al., (2025) developed a multiplex qPCR strategy compared to classical microscopy for intestinal protozoa diagnosis. Despite the high specificity and sensitivity of their qPCR assay, they reported that some microscopy-positive results were PCR-negative when detecting *Dientamoeba fragilis* and *Blastocystis* spp.

In this study, the fLAMP assay presents encouraging results for *Blastocystis* spp. diagnosis, exhibiting a sensitivity of 91.43%, specificity of 93.26%, and accuracy of 92.27% when compared to RT-PCR across 194 clinical specimens **(Table 2)**. While demonstrating strong agreement with the reference method in 179 samples, the presence of six false positives and nine false negatives warrants careful consideration. The PPV of 94.12% and NPV of 90.22% suggest a good likelihood of true results following a positive or negative fLAMP test, respectively. These performance metrics must be evaluated in the context of fLAMP’s potential advantages over RT-PCR, namely its simplicity, cost-effectiveness, and relatively earlier detection time. When comparing this assay to previously developed PCR protocols for *Blastocystis* spp. detection (Menounos et al., 2008; Stensvold et al., 2006; 2009b; Santín et al., 2011), the LAMP assay exhibits much higher sensitivity.

Like fLAMP, the cLAMP assay demonstrated a high sensitivity of 95.77% (n = 151). This indicates a strong ability to diagnose true positive *Blastocystis* spp. infections. The NPV of 95.77% further reinforces this, suggesting that a negative cLAMP result is highly reliable in ruling out infection. This is a significant strength, particularly in scenarios where minimizing false negatives is crucial to prevent the spread of *Blastocystis* spp. The discrepancy with the RT-PCR, where 136 out of 151 samples agreed, reveals the presence of FP and FN. In addition, cLAMP failed to detect *Blastocystis* spp. in three positive samples (FN). This discrepancy could be attributed to some factors that might affect the reaction sensitivity and accuracy. For instance, the potential existence of LAMP reaction inhibitors, the pathogen load in the copro-DNA, the quality of target DNA, etc. Further experiments are needed to understand the factors that led to this discrepancy. However, neither the microscopy nor the nested PCR (in addition to PCR and RT-PCR) is a perfect gold standard method for the diagnosis of pathogens in copro-DNA samples, such as *P. falciparum* (Zhang et al., 2022). This discrepancy in our results between LAMP and RT-PCR may be explained by the fact that RT-PCR is not a perfect standard, with misdiagnosis, as it sometimes produces mis-amplification in our experiment. On the other hand, we need to stress that the sensitivity of the RT-PCR assay should also be questioned, particularly when DNA samples of a lower quality and quantity were used. The RT-PCR failed to amplify the target gene in low *Blastocystis* spp. loaded samples, while some of those low-copy-number DNA samples were detected by the cLAMP and fLAMP assays. Moreover, over the Ct value of 35, the standard RT-PCR primers are prone to produce misamplifications, as shown in **Figure 1A (**sample no:150). This lowers the accuracy of RT-PCR performance. This study utilized the most common RT-PCR primer sequences that have been used in *Blastocystis* spp. diagnosis in the literature (Poirier 2021; Nguyen et al. 2023). Nevertheless, this primer composition is prone to producing misamplification and decreases the assay’s sensitivity. It is concluded that more specific primer sequences may help to eliminate misamplifications in the reactions containing low concentrations of *Blastocystis* spp. DNA.

The cLAMP assay offers significant advantages over RT-PCR results, primarily stemming from its potential for visual interpretation. Unlike RT-PCR, which typically requires specialized and often expensive instruments, cLAMP can be designed to produce a color change that is visible to the naked eye. This eliminates the need for sophisticated plate readers and complex data analysis software, making it particularly advantageous in resource-limited settings, field diagnostics, or for rapid screening where immediate results are paramount. This visual readout simplifies the testing process, reduces the reliance on trained personnel for interpretation, and can potentially lower the overall cost per test by removing the instrumentation barrier. Moreover, cLAMP’s rapid detection time, combined with its ease of visual interpretation, provides a compelling alternative for qualitative *Blastocystis* spp. detection, particularly when speed and accessibility are critical factors, outweighs the need for precise quantification. The direct, visual nature of cLAMP results can also facilitate immediate action and decision-making at the PoC testing.

The data provided clearly indicate a significant advantage in the mean time-to-detection threshold (TT) for fLAMP and cLAMP **(Figure 5)**. The mean TT for the cLAMP was 29.2 minutes. This is notably faster than the reported average of 36.2 minutes for the initial results of RT-PCR. It should be noted that the TT for both RT-PCR and fLAMP is based on the threshold cycle (Ct) value, showing the initial time point where the amplification curve started to ramp. However, the total reaction time, which includes the amplification and melting curve analysis, takes about 90 minutes. Furthermore, the fLAMP assay demonstrated an even faster average detection time of just 24.0 minutes. The reaction of fLAMP and cLAMP is complete in less than an hour (55 minutes). This substantial reduction in assay duration positions both fLAMP and cLAMP as potentially robust alternatives to RT-PCR, particularly in PoC settings where rapid results are crucial for timely clinical decision-making and effective disease management.

The necessity of performing a DNA extraction step on stool specimens is a drawback of this *Blastocystis* spp. detection method, as it increases the overall turnaround time for diagnosis. Furthermore, subtyping *Blastocystis* spp. necessitates a complementary molecular technique (e.g., RT-PCR followed by sequencing). It is also not possible to directly sequence LAMP products because they form various loop and dumbbell-like structures of different sizes.

## CONCLUSION

Fluorometric loop-mediated isothermal amplification (fLAMP) and colorimetric LAMP (cLAMP) assays were both shown to be rapid and sensitive diagnostic tools for *Blastocystis* spp. Both methods achieved detection within a clinically relevant timeframe of 15–50 minutes. Notably, the cLAMP assay provides a significant advantage due to its simple, naked-eye color change for result interpretation, which eliminates the need for expensive, sophisticated instrumentation. This makes it particularly suitable for resource-limited settings. Both assays demonstrated promising performance metrics, with fLAMP exhibiting 91.43% sensitivity and 92.27% accuracy, while cLAMP showed 95.77% sensitivity and 90.07% accuracy. Their capacity for femtogram-level detection makes these assays valuable for analyzing samples with a low copy number of *Blastocystis* spp. DNA. In conclusion, the high analytical and diagnostic sensitivity and rapid detection time of both LAMP assays make them valuable for point-of-care (POC) testing in local screening centers and hospitals. Further studies could focus on identifying *Blastocystis* subtypes for a more precise diagnostic approach.

## Data Availability

All data produced in the present study are available upon reasonable request to the authors

## Declarations

## Ethical Approval

The Institutional Review Board (IRB) at Imam Abdulrahman bin Faisal University (IAU) approved the study with IRB number IRB-2024-13-530. The de-identified leftover samples after completion of diagnostic tests were used; hence, this study requires no consent as per institutional ethics committee regulations.

## Acknowledgement

We would like to thank the Institute for Research and Medical Consultations (IRMC) at Imam Abdulrahman bin Faisal University (IAU) for their financial and technical support. We would also like to thank Ms. Moneerah Alsaeed for her technical support.

## Conflict of interest

The authors declare no competing interests.

## Funding

This study was financially supported by the Institute for Research and Medical Consultations (IRMC) at Imam Abdulrahman bin Faisal University (IAU).

## Availability of data and materials

Not applicable

## Author contributions

HT and AAE conceptualized and designed the study. RZ and HT carried out experiments. AAE extracted and provided DNA samples. RZ and HT wrote the manuscript. All authors read and commented on the manuscript.

## Notes

### Competing Interest Statement

The authors have declared no competing interest.

### Funding Statement

This study was supported by the Institute for Research and Medical Consultations (IRMC) at Imam Abdulrahman bin Faisal University.

### Author Declarations

The Institutional Review Board (IRB) at Imam Abdulrahman bin Faisal University (IAU) approved the study with IRB number IRB-2024-13-530.

