## Supplementary material for "Development of Colorimetric and Fluorometric Loop-Mediated Isothermal Amplification (LAMP) Assays for the Point-of-Care Molecular Diagnosis of *Blastocystis* spp": Figure S1, Figure S2, Table S1

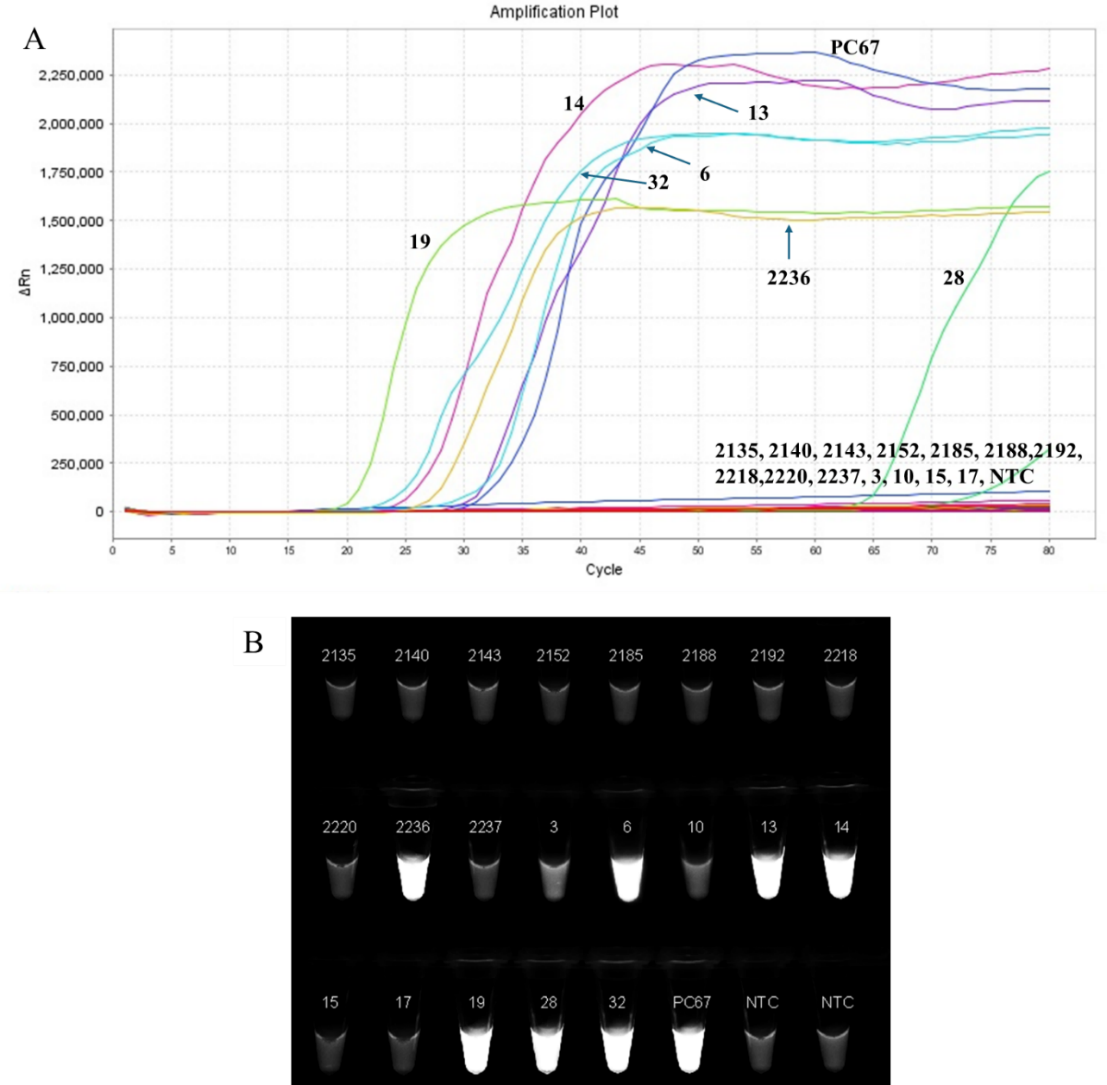

**Figure S1** Isothermal amplification of target *SSU rRNA* gene by using fluorometric LAMP. **A)** Fluorometric LAMP amplification plot. **B)** fLAMP tubes under UV transilluminator.

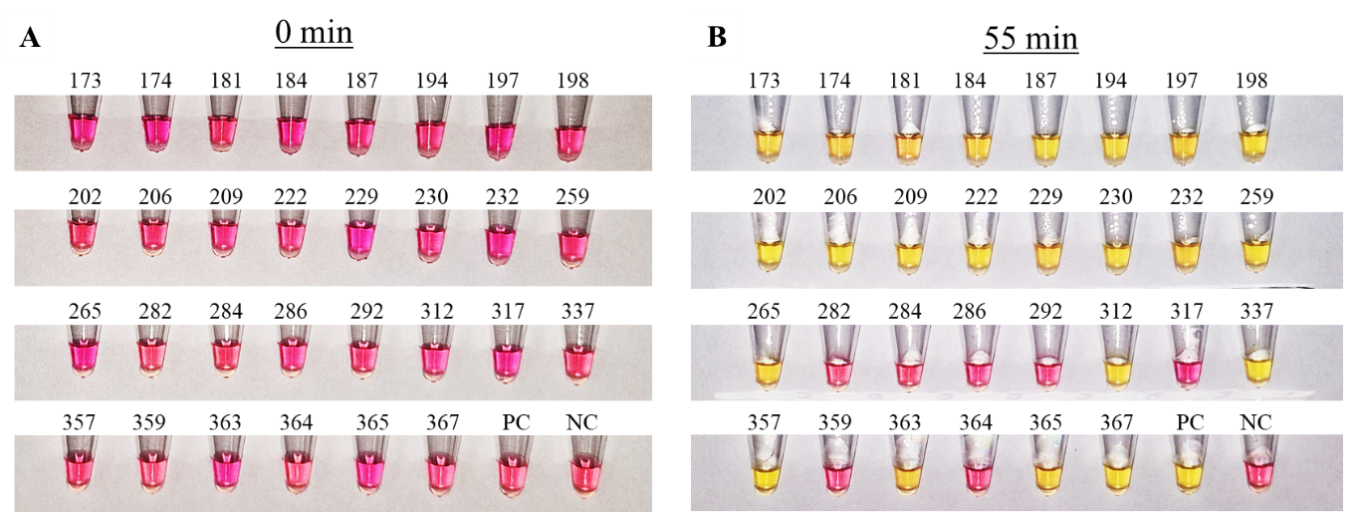

**Figure S2** Colorimetric detection of *Blastocyst* spp. Comparison of the reactions **(A)** before (0 minutes) and **(B)** after the reaction (55 minutes). This group includes 30 unknown samples with a positive control (PC) and negative control (NC) reaction tubes.

**Table S1.** The samples used to test the specificity of the assay.

| **Sample Code** | ***Cryptosporidium*** | ***Giardia*** | ***Entamoeba*** | ***Dientamoeba*** | ***H. Pylori*** | ***Blastocystisis* spp.** | |
| --- | --- | --- | --- | --- | --- | --- | --- |
|  |  |  |  |  |  | **RT-PCR** | **cLAMP** |
| 1 | - | - | + | - | na | - | - |
| 3 | - | - | + | - | na | - | - |
| 4 | - | - | + | - | na | - | - |
| 38 | - | - | + | - | na | - | - |
| 47 | - | - | + | - | na | - | - |
| 55 | - | - | - | + | na | - | - |
| 64 | + | + | + | - | na | - | - |
| 103 | + | + | - | - | na | - | - |
| 104 | + | + | - | - | na | - | - |
| S4 | na | na | na | na | + | na | - |

* na: not applicable.
